# Barriers and facilitators to the implementation of cell phone interventions to improve the use of family planning services among women in Sub-Saharan Africa: a systematic review

**DOI:** 10.1101/2022.04.24.22274232

**Authors:** Abibata Barro, Patrice Ngangue, Nestor Bationo, Dieudonné Soubeiga, Yacouba Pafadnam, Safiata S. Kaboré, Hermann Pilabré, Doulaye Traoré

## Abstract

**Background:** Mobile health (mHealth) interventions are being tested to improve contraceptive uptake in SubSaharan Africa (SSA). However, few attempts have systematically reviewed the mHealth programs aiming to improve family planning (FP) services among women in SSA. This review identifies and highlights facilitators and barriers to implementing cell phone interventions designed to target women FP services.

**Methods:** Databases including PubMed, CINAHL, Epistemonikos, Embase, and Global Health were systematically searched for studies from January 01, 2010, to December 31, 2020, to identify various mHealth interventions used to improve the use of FP services among women in SSA. Two authors independently selected eligible publications based on inclusion/exclusion criteria, assessed study quality and extracted data using a pre-defined data extraction sheet. In addition, a content analysis was conducted using a validated extraction grid with a pre-established categorization of barriers and facilitators.

**Results:** The search strategy led to a total of 8,188 potentially relevant papers, of which 16 met the inclusion criteria. The majority of included studies evaluated the impact of mHealth interventions on FP services; access (n = 9) and use of FP outcomes (n = 6). The most-reported cell phone use was for women reproductive health education, contraceptive knowledge and use. Barriers and facilitators of the use of mhealth were categorized into three main outcomes: behavioral outcomes, data collection and reporting, and health outcomes. mHealth interventions addressed barriers related to provider prejudice, stigmatization, discrimination, lack of privacy, and confidentiality. The studies also identified barriers to uptake of mHealth interventions for FP services, including decreased technological literacy and lower linguistic competency.

**Conclusion:** The review provides detailed information about the implementation of mobile phones at different healthcare system levels to improve FP services; outcomes. Barriers to uptake mHealth interventions must be adequately addressed to increase the potential use of mobile phones to improve access to sexual reproductive health awareness and family planning services.

**Systematic review registration:** PROSPERO CRD42020220669 (December 14, 2020)

## INTRODUCTION

Improving access to family planning (FP) information can be a strategy to improve service utilization and prevent adverse pregnancy outcomes and unsafe abortions among adult women of reproductive age, reduce the risk of maternal mortality, and promote the realization of reproductive rights.^1^ Indeed, lack of knowledge about appropriate FP methods, fear of side effects, myths and misconceptions about contraception are well-known barriers to FP services.^2^ In addition, lack of provider time and FP counselling opportunities may contribute to low FP service utilization in well-attended maternal child health (MCH) and clinics.^1, 3^ Thus, new approaches to providing comprehensive, client-centered FP counselling that addresses individual and structural barriers ^4^ are essential to increasing reproductive-age FP service utilization among adult women.

eHealth is defined as health care practices supported by electronic processes and communications and includes (mHealth), which in turn is often defined as the practice of medicine and public health supported by mobile devices such as cell phones, patient monitoring devices, personal digital assistants and other wireless devices.^5,6^ mHealth technologies have shown benefits in a variety of Sexual and Reproductive Health (SRH) settings in resource-limited settings.^7,8^ Many low- and middle-resource countries with limited Internet or publishing resources have achieved a significant cell phone infiltration rate.^4^ According to the 2018 International Telecommunication Union (ITU) report, the total number of cell phone subscribers worldwide has reached 5 billion people.^9^ This number is likely to increase and exceed the world’s population in the coming years due to the growing dependence on cell phone technology and cell phones’ falling costs.^10-12^ In low-income countries populations (LICPs), the cell phone infiltration rate has been over 90%.^13^ A survey was conducted in 24 developing countries to assess cell phone ownership. The survey report found that more than half of the countries’ populations have a cell phone. In addition, a median of 78% of cell phone users in 24 countries uses short messages (SMS), making it the most popular method of communication.^14^ Furthermore, cell phone owners were more likely to use modern contraceptives compared to non-owners;^15^ mHealth technology can help overcome some barriers, including provider bias, stigma, discrimination, fear of rejection, lack of privacy and confidentiality, an embarrassment in seeking SRH education and services on sensitive topics, cost issues, and transportation challenges by providing safe, accurate, cost-effective, timely and tailored FP services to women.^4^ SRH programs in SSA using mHealth have focused primarily on behavior change communication, sharing family planning knowledge through an SMS,^4,16^ either within a general population or for a target audience of adolescents,^17-19^ resulting in increased use of these services.

As a platform, mHealth has offered educational information about sexual and reproductive health and family planning service providers.^20^ Additionally, mHealth offers individuals fewer logistical barriers because they can quickly, conveniently, and confidentially seek information about family planning and related resources instead of going to a clinic or seeing a health care provider obtain the same information.^21^ Several interventions have been implemented to assess whether mHealth technologies could be used to help reduce unmet contraceptive needs in SubSaharan Africa by attempting to increase the use of family planning services.^1,18,22-35^

In recent years, there has been an increased amount of research on the potential of mHealth for women’s SRH services in SSA. However, little evidence exists on cell phone interventions to improve women’s use of FP services in SSA. Previous studies have attempted to review mHealth interventions in Low-Income Countries to address maternal health. Three hundred and seventy papers were found in the literature search. They assessed the full text of 57 studies and included 19 in the review. Studies demonstrated promise for mHealth in maternal health; however, much of the evidence came from low- and moderate-quality studies.^36^ A systematic review by Feroz and colleagues assessed the use of mobile phones to improve young people SRH in low and middle□income countries. The review provides detailed information about mobile phones’ implementation at different healthcare system levels for improving young people’s SRH outcomes, but there is a lack of literature from women of reproductive age in general.^37^ Another review by Gahungu & al. summarized 79 studies in a systematic review of the literature, which focused on unmet needs for modern FP methods among postpartum women in SSA.^38^ Unmet needs in postpartum FP in women from SSA were associated with health-system and socio-demographic determinants. However, there is a need to improve awareness of modern contraceptive methods through effective interventions.^38^ These three reviews included evidence regarding the use of mHealth to improve young people’s SRH and women in postpartum. However, very little is known regarding the potential barriers and facilitators for the uptake of mobile phone interventions to improve women’s use of family planning services. This systematic review aimed to highlight potential barriers and facilitators for the uptake of cell phone interventions to improve FP services among women in SSA. Thus, developing and implementing a cell phone FP intervention for adult women of reproductive age will inform the future use of mHealth to deliver FP programs in SSA.

We present the following article in accordance with the PRISMA reporting checklist (see supplementary file).

## METHODS

This systematic review examines barriers and facilitators to implementing mobile phone interventions to improve FP services use among women in SSA. Additionally, this review will help the research community make decisions regarding new methodologies and mobile phone interventions to encourage women of childbearing age to seek family planning information and services.

The review protocol was registered in the International Prospective Register for Systematic Reviews (PROSPERO) CRD42020220669 on December 14, 2020.

### Information sources and search strategy

An electronic systematic literature search was carried out to explore mobile phone technology’s role in improving women use of family planning services, particularly in SSA. Although there are many databases on this pertinent topic, we searched five electronic databases, including PubMed, CINAHL, Epistemonikos, Embase, and Global Health, as they are considered databases for systematic reviews. These databases were explored using a detailed search strategy. Additionally, grey literature (non-published, internal, or non-reviewed papers, repositories) was also explored as an important source for mobile phone evaluations. The reference list of included records was also appraised to identify relevant articles. The search strategy included five categories of keywords: mHealth, Women, SubSaharan Africa, Facilitators and Barriers. These keywords should appear in conjunction with the title or abstract of the article. For example, to refer to m-Health, articles either had to include the term “mhealth” (and its alternative formulations) or include both the term “health” and one of the following search terms or their variants: mobile phone, smartphone, mobile application, mobile app, cellular phone, mobile device, mobile technology, SMS, or text message. To refer to women, we used the following search terms: female, young adult, women of childbearing age, women of reproductive age. To refer to SubSaharan Africa, we used the following search terms: Africa South of the Sahara, Sub-Saharan Africa, Africa Central, Africa Eastern, Africa Southern, Africa Western. Finally, we searched the following themes related to barriers: availability of electricity, availability of mobile phone, phone cost, literacy, inadequate information on implementation costs; and facilitators: use of mobile phone, Adequate information on implementation, political stability and support, social acceptability.

Duplicate citations across databases were identified and excluded using Zotero, and a manual revision was done for verification. If a study was reported in more than one publication and presented the same data, we only included the most recent publication. However, all were included if new data were presented in multiple publications describing the same study.

### Eligibility criteria

We included studies with an abstract in English or French. The studies had to be based on an empirical design, including qualitative, quantitative or mixed-methods. The articles should clearly state the data collection process, research methods, and measurement tools. We excluded publications presenting editorials, comments, position papers, and unstructured observations from this review. We included conference proceedings as long as they presented all relevant data. Studies provided data on women’s barriers and facilitators to use mHealth in their results or discussion sections. We excluded studies that focused on studies involving groups of women, men, and girls under the age of 15□years and over 45□years.

### Studies selection

A citation management system (Zotero) was used to manage the records exported from electronic databases (40). A pre-defined screening form was developed to ensure the reliability of screening articles among the two reviewers (BA and KSS), and pilot testing was conducted per the eligibility criteria. After reviewing the studies, both reviewers (BA and KSS) described outcome measures to verify the articles’ relevance. Each reviewer provided strong justifications for excluding studies. In a consensus meeting, a third reviewer (PN) resolved disagreements between the two reviewers. The third reviewer was consulted to decide whether the study meets the eligibility criteria for inclusion.

Titles, abstract first screened all studies, and full text to progressively eliminate studies not meeting the inclusion criteria. Database searches identified a total of 8188 studies initially. After de-duplication, 8020, potentially relevant titles were included for title or abstract screening. Next, full texts of the remaining 52 studies were reviewed to determine if they fulfilled the inclusion criteria. Finally, 16 studies were selected and used for this review.^1,18,22-35^ The Preferred Reporting Items for Systematic Reviews and Metaanalyses (PRISMA) flow diagram was used to report the study selection process (Fig. 1).

**Figure 1:**
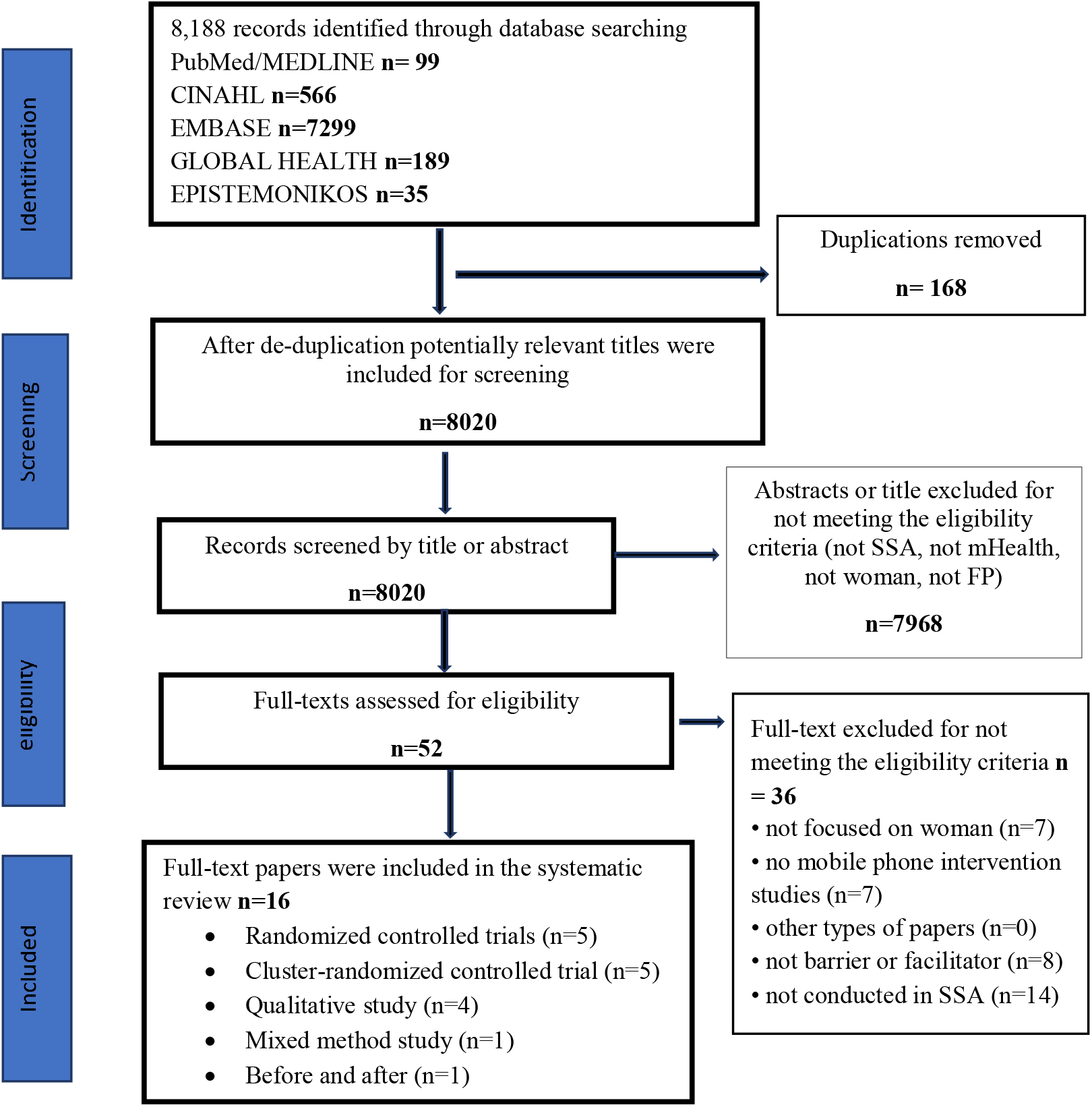
PRISMA flow diagram for database search of studies

### Data extraction

Two authors independently extracted data from each study that fulfilled the inclusion criteria using a standard form. Study characteristics (name of the first author, year of publication, the country in which the study was conducted, study design, sampling approach, participating characteristics) and key findings related to factors associated with the use of FP services by women were extracted. Any factors associated with using FP services were listed, and the results of multivariate statistical tests for association (odds ratio) were noted. For studies where a multivariate statistical test was not done, the results of bivariate analyses were noted. When the measure of association in multivariate analysis was not significant and not reported by authors, the factor was not considered in the synthesis.

### Quality assessment of included studies

We used the mixed methods appraisal tool (MMAT) 2018 version to assess the methodological quality of the included studies. The MMAT is a critical appraisal tool designed to appraise qualitative, quantitative and mixed methods studies. It allows the appraisal of five methodological quality categories: qualitative research, randomized controlled trials, non-randomized studies, quantitative descriptive studies, and mixed methods studies.^41^ The tool is divided into two parts. First, the tool was suited for this review as it was specifically developed for quality appraisal in systematic reviews involving qualitative, quantitative and mixed methods designs. Qualitative and quantitative sections have four criteria each, and studies are scored by dividing the number of criteria met by four to arrive at a value ranging from 25 to 100%. For mixed-method studies, we adapted the MMAT by assessing each segment separately and then selecting the lowest quality rating. Articles were not excluded based on the MMAT score; the purpose was to examine and gain insight into the rigor of existing research in this field.

### Data synthesis and analysis

The reviewers identified sections of the publications that presented a relevant barrier or facilitator to using mHealth for the women to improve their use of FP services. In addition, the authors coded each facilitator and barrier according to the Consolidated Framework for Implementation Research (CFIR), a conceptual framework created to guide the systematic assessment of factors that influence the implementation and effectiveness of interventions.^42^ The CFIR codebook describes five broad domains and 39 constructs in much detail.

Two authors collaborated to produce one consolidated document containing all relevant codes by resolving disagreements between their original data extraction documents. Then, a third author was brought in to resolve any conflicts between documents. The two authors agreed roughly 70% about extracting barriers and facilitators for each article and 75% about the coding. Finally, all barriers and facilitators were grouped inductively to facilitate the creation of broad themes.

## RESULTS

### Included Studies

Our search identified 8188 papers. We removed 168 duplicates and screened 8020 titles and abstracts, of which 7968 were excluded after initial screening. Full texts of 52 studies were assessed; 36 were excluded, and 16 were included in this review. The study selection flow diagram is presented in Figure 1.

### Characteristics of included studies

Of the 16 papers, ten (10) described programs in Kenya, ^1,17,23–26,29,31,32,34^ 2 in Malawi,^33,35^ and 1 in Ghana (18), Uganda,^27^ Nigeria,^28^ Tanzania.^30^ Each paper reported different outcomes and/or study periods. Nine papers reported interventions targeting couples, four on interventions targeting Postpartum women, three targeted users; one sex worker; and one on interventions targeting students. Three papers were graded low quality, four (4) moderate, and nine (9) high. Eight papers described reported on randomized controlled trials (RCTs), whereas five reported on a cross-sectional qualitative study. One study used a before and after design with no control group, two studies used a pilot study. Sample sizes ranged from 12 to 7,397, limiting comparability.

The largest proportion of papers was classified as high quality. Of the nine (9) high-quality papers, four (4) reported on randomized controlled trials (RCTs), two (2) reported on cluster-randomized control trials, one reported before and after studies, and two reported on Cross-sectional data. The four moderate-quality papers reported on a qualitative study, and 1 was a Pilot study on the general public. The remaining three low-quality papers: one reported on RCTs, one pilot study (quantitative and qualitative data) and one reported on Cluster-randomized controlled trial. Characteristics of included studies are presented in table 1.

**Table 1.** Characteristics of articles included in the systematic review N=16

| Authors | Country | Target of Intervention | Goal | Duration | Intervention | Design | Quality |
| --- | --- | --- | --- | --- | --- | --- | --- |
| Johnson et al, 2017 | Kenya | Consumers | Knowledge and use of contraception | September 2013–May 2014 | The m4RH service: via text message - 13,629 Consumers | RCT | High |
| Jones and al, 2020 | Kenya | Women | Care-seeking behavior and uptake of family planning | November 2017 and concluded in March | Postpartum Checklist (PPC) messages; General Postnatal Care messages<br>Family Planning messages | RCT with four study arms. | High |
| Rubee Dev et al, 2019 | Kenya | Postpartum women and FP providers | Acceptability and feasibility of the iMACC mobile decision aid | 6 week | Interactive Mobile Application for Contraceptive Choice [iMACC]: self-administration ; combines images and text in a heuristic approach | A cross-sectional qualitative study | High |
| Lee et al, 2019 | Kenya | Female respondents | Exposure to family planning messages through mHealth and contraceptive knowledge and use | May and September 2015. | mHealth exposure was defined as having received family planning messages through a mobilephone via text or e-mail in the last 12 months | Before and after with no control group | High |
| Harrington et al, 2019 | Kenya | Postpartum women | 2-way short message service (SMS) with a nurse on postpartum contraceptive | July 19 and December 6, 2016 | Health education : contraceptive information via text messaging (Mobile WACH XY) | Unblinded RCT | High |
| Rokicki et al, 2017 | Ghana | Female students | Text-messaging programs to improve reproductive health | 3-month follow-up; 15-month follow-up | An interactive mobile phone quiz on reproductive health knowledge at 3 and 15 months;<br>Unidirectional intervention sent participants text messages with reproductive health information;<br>Interactive intervention engaged adolescents in text-messaging reproductive health quizzes | Cluster-RCT | High |
| Ampt et al, 2020 | Kenya | Sex workers | The mHealth to reduce the incidence of unintended pregnancy | Sept 14, 2016, and May 16, 2017 | WHISPER message content focused on promotion of contraception | Cluster-RCT | Low |
| Nuwamanya et al, 2020 | Uganda | Students | a mobile phone application (APP) to increase access to SRH information, goods, and services | Oct 23rd, 2018, to 13th November 2018 | Participants granted access to a MPA over a period of six months. | RCT | Low |
| Babalola et al, 2019 | Nigeria | Women | Exposure to the digital tool on contraceptive ideation and use | March 7, 2017, to June 5, 2017 | The Smart Client digital health tool was designed to be delivered via mobile phone and included 17 prerecorded calls : 1 welcome call, 13 regular program calls, and 3 quiz calls interspersed. | A cluster-randomized control trial | High |
| Harrington et al, 2019 | Kenya | men and Women | SMS to facilitate postpartum FP counseling | April to June 2016 | Theoretical framework for SMS development : SMS messages based on behavioral theory and experience | Focus group discussions (FGD) among men (n = 35) and among pregnant/postpartum women (n = 15) | Moderate |
| L'Engle et al, 2013 | Tanzania | General public | The feasibility, reach and potential behavioral impact of providing automated family planning information | September 2010 through June 2011 | Every m4RH user logged by the system was sent a series of four questions via text message. | Pilot study (2870 unique users accessed m4RH) | Moderate |
| Shelus et al, 2017 | Kenya | Female app users | CycleBeads app brings : experience of the new users to family planning | May and September 2015 | The CycleBeads app, a digital platform to support women using the Standard Days Method, is the first mHealth app marketed in Kenya to offer an evidence-based family planning method exclusively via mobile phone. | A three-month pilot study (quantitative and qualitative data from 185 female app users.) | Low |
| Unger et al, 2018 | Kenya | Postpartum women | Short message service (SMS) communication on contraceptive use | August 2013 and April 2014 | Women were to receive 1-way SMS versus 2-way SMS with a nurse versus control. | 3-arm, unblinded RCT | High |
| Hu et al, 2020 | low-middle-income countries | Mothers | Received SMS-based family planning communication and use of modern contraception |  | Mothers receiving SMS regarding family planning, aswell as the association between receiving SMS with modern contraceptive use, and utilisation of essential maternal health services | Cross-sectional data 94,675 mothers (15–49 years) | High |
| Vahdat et al, 2013 | Kenya | Users | The m4RH access and potential behavioral impact of providing contraception information | January 2010 to June 2011 | Automatic logging of all queries to the m4RH system ; demographic and behavior change questions and telephone interviews with a subset of m4RH users. | The structured interview | Moderate |
| Laidlaw et al, 2017 | Malawi | Stage 1: representing men, women, leadership, elderly and male and female youth;<br>Stage 2: male adults, female<br>adults, male<br>youth and<br>female youth | Implementing an mHealth intervention | Stage 1: secondary analysis of village profiles in 2013/14.<br>Stage 2: intervention development in 2015. | mHealth education intervention, an mHealth messaging service, which will provide health information to the community | Two stage qualitative study | Moderate |

### Overview of mHealth using factors

In total, 64 elements were identified as barriers to or facilitators for mHealth use and were classified in the CFIR domains and constructs from the extraction grid. 41 (64.06%) of these elements were classified as facilitators for mHealth adoption and 23 (35.94%) as barriers. The reported frequency of the barriers and facilitators and their alignment to the CFIR constructs are shown in Table 2.

**Table 2.** Frequency of barriers and facilitators identified from included items.

| **CFIR domains and constructs** | **Barriers (N)** | **Facilitators (N)** | **Total** |
| --- | --- | --- | --- |
| **Domain 4: Individuals involved in the implementation** | **15** | **16** | **31** |
| Awareness of the existence of mHealth | 1 | 1 | 2 |
| Familiarity, ability with mHealth | 4 | 1 | 5 |
| Familiarity with technologies in general | 1 |  | 1 |
| Risk-benefit assessment (perception) | 2 |  | 2 |
| Autonomy | 2 | 1 | 3 |
| Outcome expectancy | 3 | 9 | 12 |
| Agreement with mHealth (Welcoming/resistant) | 1 | 2 | 3 |
| Belief in one's competence to use mHealth |  | 1 | 1 |
| Voluntary ownership |  | 1 | 1 |
| Experience | 1 |  | 1 |
| **Domain 2: Outer Setting** | **3** | **8** | **11** |
| Womens' attitudes and preferences towards mHealth |  | 2 | 2 |
| Women and health professional interaction |  | 5 | 5 |
| Applicability to the characteristics of women | 2 |  | 2 |
| Other factors associated with women | 1 | 1 | 2 |
| **Domain 1: Characteristics of the Intervention** | **5** | **17** | **22** |
| Design and technical concerns | 2 |  | 2 |
| Perceived usefulness |  | 10 | 10 |
| Perceived ease of use | 1 |  | 1 |
| Satisfaction about the content available (completeness) |  | 4 | 4 |
| Content appropriate for the users (relevance) |  | 3 | 3 |
| Cost issues | 1 |  | 1 |
| Cell phone accuracy | 1 |  | 1 |
| **Total** | **23** | **41** | **64** |

#### Individuals involved in the implementation

Individual factors represented 31 (48.44%) of the extracted elements. There were twice as many facilitators as barriers in this category (41 and 23, respectively). The most common factor identified was outcome expectancy (n=12).^1,18,22–24,26–29,32,33^ Most women valued mHealth and described it as useful in helping to dispel myths and misconceptions, setting realistic expectations about potential side effects of FP and maintaining confidentiality. Indeed, mobile health could be considered a cost-effective tool for improve+ng family planning knowledge.^1,17,18,23,26–29,32^ However, some pointed out that mHealth alone did not improve contraceptive knowledge and modern contraception among women.^17,24,33^ Familiarity and ability with mHealth are considerable skill limitations in low-literacy settings, especially regarding recruitment and induction of women (n=4).^17,24,28,43^ Awareness of the existence of mHealth (n=2)^26,43^ was perceived either as a facilitator or a barrier, awareness dependent on other factors, such as familiarity, ability with mHealth and technologies in general.

Autonomy, agreement with mHealth (Welcoming/resistant) were also mentioned three times.^1,24,26,28^ In addition, one comment underlined that future adaptations of the tool should address the limitations connected with the number and length of program calls.^28^ Finally, familiarity with technologies in general (1), voluntary ownerships (1), experience (1), and beliefs in one’s competence to use mHealth ^28^ were other factors identified in this category.

#### Outer Setting

Outer Setting represented eleven of the elements identified in the review (17.19%). Eight of the factors extracted were facilitators, and three were barriers. Women and health professional interaction (n=5)^18,25,28,29,32^ were underlined more often than other factors. Professionals believed that mHealth, especially in the case of smartphones, SMS dialogue with a nurse about FP could reduce misperceptions and stimulate communication within couples, thereby improving contraceptive access and continuation.^18,25,28,29,32^ Other factors related to women were applicability to the patients’ characteristics^17,28^, women’s attitudes and preferences towards mHealth ^23,35^ and other factors associated with women.^24,35^

#### Characteristics of the Intervention

A total of 22 elements (34.37%) pertain to the category “Characteristics of the Intervention”, with five of them identified as barriers and 17 as facilitators. The most recurrent using factor was perceived usefulness, with ten extracted elements.^1,17,18,26–31,33^ It was seen as a facilitator for mHealth adoption. Perceived usefulness is defined as an individual’s perception that the utilization of a particular mobile device will be advantageous in an organizational setting over a current practice.^44^ Satisfaction about content available (completeness) was another frequently mentioned factor (n=4).^1,23,28,32^ Satisfaction about the content available is defined by personalized, complete, relevant, easy to understand and secure information content.^45^ Therefore, it was important for the women to perceive messages’ usefulness and content completeness in their living environment; otherwise, there would be less incentive to use them. Content appropriate for the users (relevance) was also mentioned on three occasions^24,32,34^ and was perceived mainly as facilitators.

Other factors related to mHealth characteristics were perceived ease of use,^43^ cost issues,^24^ and cell phone accuracy ^24^ (all of them extracted one time). Cost issues, perceived ease of use and cell phone accuracy were seen exclusively as barriers to the use of mHealth. Indeed, women were worried about the cost of mobile technology and smartphone applications were perceived as barriers to mHealth use. Additionally, women were worried about literacy and lack of familiarity with smartphones or tablets and suggested the inclusion of interactive multimedia such as audio or videos to optimize the tool’s effectiveness. Design and technical concerns were other factors identified in this category.^1,28^

## DISCUSSION

### Summary of findings

This systematic review is the first to assess factors influencing the implementation of mhealth interventions toward increasing the use of family planning services in SSA. Other systematic reviews have examined mHealth in family planning interventions, but few included studies from SubSaharan Africa. Additionally, three of the 16 studies in the present review were not assessed in previous systematic reviews. Findings from the current systematic review reveal new information about the role that mHealth has in improving the use of family planning services in SubSaharan Africa. The mHealth solutions identified in this systematic review mainly aimed to improve FP related SRH education, services and behavioral outcomes for women.

Out of the 16 studies included, three reported improvements in family planning behavioral outcomes among women who received the intervention compared with controls.^23,26,28^ Concerning mHealth, two of the three studies used text messages,^23,26^ while the other study used voice messages and telephone counseling, which included information about the nearest family planning service provider.^28^ Thus, one common trait that the three studies shared was interactive communication to deliver tailored intervention content to participants. Other commonalities were the use of motivational messages ^23,26^ and the involvement of a male partner in the intervention.^28^

Seven of the 16 studies found improvements in family planning outcomes, the full extent that cell phones contributed to improvements in the use of family planning services among participants cannot be determined. Certain types of mHealth features may be more advantageous to effect change in the use of contraceptives. For example, Lee and colleagues found that mHealth alone was limited in improving contraceptive knowledge and use in their study conducted in Kenya. However, mHealth led to intended outcomes when used with four other channels.^24^ When to Hu et al., receiving SMS on family planning does not affect modern contraceptive use. However, a significant increase in the odds of availing health facility delivery services among those receiving SMS about family planning is notifie.^33^ This review found that text message-based health interventions are feasible and acceptable for improving women’s reproductive health information and access to contraceptives.^17,25,27^

The categorization of the studies in various mHealth applications provided the understanding that the strongest evidence exists on client education and behavior change communication mHealth application. These findings are in concordance with the other reviews, suggesting that mobile phone approaches, including texting, have been explored much by various studies. It provides a feasible and potential efficacious medium for increasing reproductive and sexual health education.^18,31,34,43^

### Comparison with Existing Literature

Based on our analysis, the most reported use of cell phones was health outcomes, including women reproductive health education, contraceptive knowledge and use,^17,23–25,27,29,32,33^ followed by data collection and reporting,^18,30,31,34,35,43^ and behavioral outcomes.^23,26,28^ The grouping of cell phone and FP studies by outcome provided the understanding that the strongest evidence exists on women’s reproductive health education, contraceptive knowledge and use. These findings are in concordance with the other reviews, suggesting that cell phone approaches, including texting, have been explored much by various studies. It provides a feasible and potentially efficacious medium for increasing education of family planning and sexual health levels.^37,46^ However, these studies are primarily conducted in developed countries than in SSA. Thus, a complete understanding of cell phones’ role in improving women’s use of family planning services is required to strengthen the evidence base in SubSaharan Africa. More studies are needed to refine the current work with a larger body of evidence and establish how best to integrate it with the published existing framework.

The International Conference on Population and Development set men’s involvement in family planning as a priority area.^47^ Harrington and coll.^29^ conducted four focus groups (FGD) among men (n = 35) and two among pregnant/postpartum women (n = 15) in western Kenya. This component may have contributed to improving FP decision-making and couple communication. For example, Tao and coll.^48^ found that the male partner’s involvement in family planning decision making improved family planning knowledge and contraceptive continuation. Prior research suggests that male partners’ involvement is advantageous for family planning and the uptake of contraceptive methods. However, future research is warranted to assess whether the type of male partners differs (e.g., sexual/romantic relationship, family, friend) and the amount and frequency of their involvement toward achieving these outcomes.

Two studies included in this review reported using a behavioral change theory.^25,43^ One used the Integrated Behavioral Model,^43^ and the other study used the Theory of Planned Behavior.^25^ They are similar derivative theories of general behavioral prediction, with the most important determinant being motivation or intention as the interventions targeted. A systematic review by Cho and coll.^49^ examined the use of theories in mHealth behavior change interventions conducted in the LMICs and found that about one-third (5 of 14) of their included studies were based on a behavioral change theory. Well-tested behavioral change theories help guide family planning interventions and programs.^49,50^ However, cell phone effectiveness in family planning interventions in SubSaharan Africa remains inconclusive. Future research using behavioral change theory for contraception uptake is warranted and needed to help identify which intervention components (cell phone and behavior change) work best for family planning.

### Importance for research and practice

The review has provided an understanding of how cell phone solutions targeting women help address issues of ‘provider’s prejudice, stigmatization, lack privacy and confidentiality, cost prohibitions, and transportation challenges’.^30,34,43^ Simultaneously, the review has highlighted the barriers to uptake mHealth solutions for FP services, including poor technological literacy, insufficient network coverage, lower linguistic competency, high cost of service, and socio-cultural beliefs and expectations not favoring the use of mHealth.^18,43^ Additionally, larger-scale and more rigorous studies are needed to assess external validity across SSA settings to guide health-sector resource investments into these technologies. Finally, future research should explore new areas of application of mHealth interventions, such as healthcare providers training, supervision and quality improvement for health workers. Despite these research needs, mHealth has significant potential to alter the landscape for using FP services in SSA and is worthy of attention and support. This opens a window to look at the issue from a broader perspective to explore the most important technology implementation challenges.

### Limitations

This systematic review has its strengths and weaknesses. To ensure a comprehensive search strategy, we used a literature search strategy adding specific terms for the three components we were interested in studying (mobile phone and women and SubSaharan Africa) and used clear inclusion and exclusion criteria. In addition, although we included only papers published in peer-reviewed journals to improve the review’s quality, this may have resulted in the omission of outside reports from nonprofit organizations, white or grey literature, or papers published in technology journals. Another limitation is that we only included papers published in English and French. Finally, it is worth noting that there is program overlap among some of the reports included in this review.

Also, we used a theoretical framework (CFIR) for classifying elements identified as barriers and facilitators to mHealth use from the studies included in this review. As such, we relabelled some of the original factors in order for them to fit within our conceptual framework. We acknowledge that the use of other theoretical frameworks or models could have uncovered other dimensions of the use of mHealth. However, we think that the framework used is comprehensive and well suited to present use factors of mHealth perceived by women of reproductive age as it is based on extensive theoretical and empirical research.

## CONCLUSION

The review provides insights for the research community and public health professionals in making decisions regarding innovative, engaging and effective mobile phone interventions to improve family planning services outcomes among women. The findings from this systematic review provide a common ground, making it possible to understand better the challenges and opportunities related to mHealth utilization by women of reproductive age. While some of the barriers and facilitators to mHealth use are similar to those identified in systematic reviews about other ICT applications, this review has enabled us to identify factors specific to mHealth.

## Data Availability

All data produced in the present work are contained in the manuscript

## Supplementary Information

**Additional File 1**: The summary of included studies on mHealth interventions to improve the use of family planning services among women

## Acknowledgements

Funding: The author(s) received no financial support for this article’s research, authorship, and/or publication.

## Authors’ contributions

BA conceived and designed the study. BA drafted the manuscript and is the guarantor of the systematic review. BA, KSS, PN developed the search strings, searched the studies, selected the studies, extracted data, and synthesized. BA, KSS, PN, BN, PY extensively reviewed the manuscript. All authors read, provided feedback, and approved the final version of the manuscript.

## Availability of data and materials

Not applicable.

## Ethics approval and consent to participate

Not applicable.

## Consent for publication

Not applicable.

